# Precision Medicine Approach to Alzheimer’s Disease: Successful Proof-of-Concept Trial

**DOI:** 10.1101/2021.05.10.21256982

**Authors:** Kat Toups, Ann Hathaway, Deborah Gordon, Henrianna Chung, Cyrus Raji, Alan Boyd, Benjamin D. Hill, Sharon Hausman-Cohen, Mouna Attarha, Won Jong Chwa, Michael Jarrett, Dale E. Bredesen

**Affiliations:** Bay Area Wellness, Walnut Creek, CA; Integrative Functional Medicine, San Rafael, CA; Northwest Memory Center, Ashland, OR; Quesgen Systems, Burlingame, CA; Department of Radiology, Washington University School of Medicine, St. Louis, MO; CNS Vital Signs, Morrisville, NC; Department of Psychology, University of South Alabama, Mobile, AL; IntellxxDNA, Austin, TX; Posit Science, San Francisco, CA; Department of Radiology, St. Louis University, St. Louis, MO; Department of Molecular and Medical Pharmacology, David Geffen School of Medicine, UCLA, Los Angeles, CA

## Abstract

**Importance:** Effective therapeutics for Alzheimer’s disease and mild cognitive impairment are needed.

**Objective:** To determine whether a precision medicine approach to Alzheimer’s disease and mild cognitive impairment, in which potential contributors to cognitive decline are identified and targeted therapeutically, is effective enough in a proof-of-concept trial to warrant a larger, randomized, controlled clinical trial.

**Rationale:** Previous clinical trials for Alzheimer’s disease have pre-determined a single treatment modality, such as a drug candidate or therapeutic procedure, that may be unrelated to the primary drivers of the neurodegenerative process. Therefore, increasing data set size to include the potential contributors to cognitive decline for each patient, and addressing the identified potential contributors, may represent a more effective therapeutic strategy.

**Hypothesis:** Alzheimer’s disease is a multi-factorial network dysfunction that results from a chronic or repeated insufficiency of support for a neuroplasticity network; thus factors that increase demand—such as infections or toxin exposure—or reduce support—such as reduced energetics or trophic support—may contribute to the neurodegenerative process. Rectifying this hypothesized network dysfunction represents a rational approach to the treatment of the cognitive decline associated with Alzheimer’s disease and mild cognitive impairment.

**Design:** Twenty-five patients with Alzheimer’s disease or mild cognitive impairment, with Montreal Cognitive Assessment (MoCA) scores of 19 or higher, were evaluated for markers of inflammation, chronic infection, dysbiosis, insulin resistance, protein glycation, vascular disease, nocturnal hypoxemia, hormone insufficiency or dysregulation, nutrient deficiency, toxin or toxicant exposure (metals, organic toxicants, and biotoxins), genetic predisposition to cognitive decline, and other biochemical parameters associated with cognitive decline. Brain magnetic resonance imaging with volumetrics was performed at baseline and study conclusion. Patients were treated for nine months with a personalized, precision medicine protocol that addressed each patient’s identified potentially contributory factors, and cognition was assessed at t = 0, 3, 6, and 9 months.

**Trial registration and IRB approval:** The clinical trial was registered at clinicaltrials.gov (NCT03883633),^1^ and approved by the Advarra IRB.

**Support for the trial:** The trial was supported by a grant from the Four Winds Foundation via Evanthea, LLC, and we are grateful to Diana Merriam and Gayle Brown for their interest, discussions, and support.

**Main Outcome Measures:** Trained external raters evaluated the study subjects with the Montreal Cognitive Assessment (MoCA), CNS Vital Signs (a computerized cognitive assessment battery), AQ-21 (a subjective scale completed by the significant other or study partner), and AQ-C change scale (a subjective scale of cognitive improvement or decline, completed by the significant other or study partner). Follow-up brain MRI with volumetrics was carried out at the completion of the trial.

**Results:** All outcome measures revealed improvement: statistically highly significant improvement in MoCA scores, CNS Vital Signs Neurocognitive Index, and AQ-C were documented. No serious adverse events were recorded.

**Conclusions and Relevance:** Based on the cognitive improvements observed in this study of patients with Alzheimer’s disease or mild cognitive impairment, a larger, randomized, controlled trial of the precision medicine therapeutic approach described herein is warranted.

## Introduction

Neurodegenerative diseases such as Alzheimer’s disease, frontotemporal dementia, and amyotrophic lateral sclerosis are without effective therapeutics. There are approximately 5.8 million people with Alzheimer’s disease in the United States, and at least one study estimates that it has become the third leading cause of death. ^2^ Unfortunately, therapeutic approaches to date have not led to sustainable improvements, and the best results from recent clinical trials have been to slow cognitive decline rather than improve cognition or halt decline.^3^

In the field of oncology, a personalized, precision medicine approach, in which the presumptive molecular drivers of the disease process are targeted therapeutically, has improved outcomes in at least some studies. ^4^ However, this strategy has not been applied successfully to neurodegenerative diseases. One complicating feature of such an application is that the etiology of Alzheimer’s disease remains controversial, with many competing theories, such as the theory that Alzheimer’s disease is “type 3 diabetes”,^5^ or is due to chronic *Herpes simplex* infection,^6^ or due to amyloid-β, ^7^ or to misfolded proteins such as tau,^8^ or prions, ^9^ among numerous other theories, none of which has led to effective treatment. However, epidemiological, pathological, toxicological, genetic, and biochemical studies have provided candidate mechanisms for the neurodegeneration associated with Alzheimer’s disease, such as neuroinflammation,^10^ insulin resistance,^11^ and reduction in trophic support.^12^

Addressing these candidate mechanisms with a precision medicine-based protocol has led to anecdotal reports of cognitive improvement in patients with Alzheimer’s disease and its forerunner, mild cognitive impairment (MCI).^13 14 15^ These anecdotal reports have provided support for the execution of a proof-of-concept trial, the results of which are presented herein.

## Methods

### Participants

Twenty-five patients with Alzheimer’s disease or mild cognitive impairment, ages 50-76, were recruited to three clinical sites: Walnut Creek, California; San Rafael, California; and Ashland, Oregon. Note that 28 patients were originally recruited, but due to COVID-19 or family circumstances, three terminated prior to the first (3-month) follow-up and were not included in the study analysis. Of the 25 patients (13 women and 12 men) who completed the study, 4 were homozygous for ApoE4, 8 were heterozygous for ApoE4, 11 were homozygous for ApoE3, and 2 were heterozygous for ApoE2 and ApoE3. Demographics are listed in Table 1.

**Table 1.** Demographics of study patients.

|  |  |
| --- | --- |
| **Age** | 50-76 |
| **Gender** |  |
| Female | 13 |
| Male | 12 |
| **ApoE alleles** |  |
| 2/3 | 2 |
| 3/3 | 11 |
| 3/4 | 8 |
| 4/4 | 4 |
| **Education** |  |
| High school | 1 |
| Some college | 2 |
| College graduate | 11 |
| Post-graduate | 11 |

**Inclusion criteria** were the following: age 45-76 years; cognitive impairment, as demonstrated by a combination of AQ-21 >5 and either MoCA of 19-26 or CNS Vital Signs <50^th^ percentile in at least two subtests or <70^th^ percentile for the Neurocognitive Index (NCI). Thus all patients had multiple areas of impairment as judged by their significant others or study partners, as well as cognitive testing indicative of MCI or dementia.

**Exclusion criteria** were the following: MoCA score <19; uncontrolled major medical illness such as seizures, cardiovascular disease, or cancer; a major psychiatric diagnosis that affected activities of daily living; ongoing psychoactive medications known to impact cognition; ongoing statin use, unless eligible to discontinue; ongoing anticoagulant therapy or history of deep vein thrombosis; MRI findings of hydrocephalus, cerebral infarct, extensive white matter disease, or intracranial neoplasm; symptomatic traumatic brain injury; lack of study partner (family member or caregiver); history of breast cancer; inability to exercise; lack of computer access; potential for pregnancy; diagnosis of a neurodegenerative disease other than Alzheimer’s (e.g., frontotemporal dementia); previous or ongoing treatment for MCI or dementia with the protocol used here or a very similar approach.

### Evaluation

Standard physical and neurological examinations were performed on each patient. Trained external raters (i.e., unaffiliated with the treatment teams) performed and assessed the Montreal Cognitive Assessment (MoCA). Computerized neuropsychological assessment batteries were performed using CNS Vital Signs, which samples multiple domains (verbal memory, visual memory, simple attention, complex attention, cognitive flexibility, executive function, processing speed, psychomotor speed, motor speed, and reaction time) as well as providing an overall Neurocognitive Index (NCI) and composite memory score. The AQ-21 (Alzheimer’s Questionnaire) is an informant-based subjective assessment with sensitivity and specificity for amnestic MCI and AD of over 90%,^16^ answered by the significant other or study partner, with scores ranging from 0 (no problems noted) to 27 (all positive responses to questions regarding impairment). A score of 5-14 suggests mild cognitive impairment, and 15-27 suggests dementia. Twenty-two of the patients in this study had AQ-21 of 6-14 and three had AQ-21 of 15-18.

**Genetic testing** was carried out using the IntellxxDNA clinical decision support tool. This allowed us to evaluate a few hundred genomic variants that can contribute to cognitive decline, including ApoE genotype, markers for hypercoagulation (e.g., Factor V Leiden), detoxification (e.g., null alleles affecting glutathione-related enzymes and other detoxification pathways), and methylation (e.g., MTHFR and MTRR), as well as a variety of other markers associated with cognitive decline such as gene variants contributing to brain hormone levels, inflammation, and nutrient transport.

**Biochemical tests and biomarkers** were performed to identify markers of insulin resistance (HOMA-IR), protein glycation (hemoglobin A1c), vascular disease (advanced lipid panel, C-reactive protein, homocysteine), systemic inflammation (C-reactive protein, fibrinogen, homocysteine), chronic infection (titers for *Herpes* family viruses (*Herpes simplex type 1, Herpes simplex type 2, Epstein-Barr virus*, and *Human herpesvirus 6*), *Borrelia, Babesia, Bartonella, Treponema pallidum, Human immunodeficiency virus*, and *Hepatitis C virus*), gastrointestinal health (stool analysis of gut pathogens, digestion, absorption, gut immune markers, and microbiome analysis), hormone dysregulation (serum estradiol, progesterone, pregnenolone, DHEA sulfate, testosterone (free and total), sex-hormone binding globulin, prostate-specific antigen (in males), free T3, free T4, reverse T3, and TSH), nutrient status (B vitamins, vitamin D, vitamin E, magnesium, zinc, copper, CoQ10, lipoic acid, omega-6:omega-3 ratio, omega-3 index), toxin or toxicant exposure (metals, organic toxicants, and biotoxins (urinary mycotoxins)), autoimmune markers (e.g., thyroid peroxidase, thyroglobulin, anti-nuclear antigen), immunoglobulins, CD57, nocturnal hypoxemia (oximetry to identify sleep apnea and upper airway resistance syndrome), and other biochemical parameters associated with cognitive decline.

**Magnetic resonance imaging** of the brain with volumetrics was performed for each patient, during initial evaluation and again at the completion of the 9-month treatment protocol. All scans at baseline and follow-up were performed on clinical 3-Tesla MRI scanners with either MPRAGE or SPGR sequences. Segmentation and quantification of the hippocampus and total gray matter were carried out computationally, as described previously ^17^. Volumetric change over the 9-month time period was calculated on a percent annualized rate of change. Additionally, each of these volume change rates was adjusted for each participant’s total head size, computed as the sum of gray matter, white matter, and cerebrospinal fluid. Rates of change in brain volumes of the hippocampus and total gray matter were then compared to historical data from the indicated references.

### Treatment

Patients were treated for nine months with a personalized, precision medicine protocol that addressed each patient’s identified potentially contributory factors, and cognition was assessed at t = 0, 3, 6, and 9 months. The goal was to identify and address the factors associated theoretically and epidemiologically (though in some cases yet to be proven causally) with Alzheimer’s-related cognitive decline: restore insulin sensitivity, improve hyperlipidemia, resolve inflammation if present (and remove the cause(s) of the inflammation), treat pathogens, optimize energetic support (oxygenation, cerebral blood flow, ketone availability, and mitochondrial function), optimize trophic support (hormones, nutrients, and trophic factors), treat autoimmunity if identified, and detoxify if toxins were identified.

The treatment team included a health coach, nutritionist, and a physical trainer, as well as the physician.

**Diet** was a plant-rich, high-fiber, mildly ketogenic diet, high in leafy greens and other non-starchy vegetables (raw and cooked), high in unsaturated fats, with a fasting period of 12-16 hours each night. Organic produce, wild-caught low-mercury fish (salmon, mackerel, anchovies, sardines, and herring), and modest consumption of pastured eggs and meats were encouraged, as well as avoidance of processed food, simple carbohydrates, gluten-containing foods, and dairy. Blood ketone levels were monitored with fingerstick ketone meters, with a goal of 1.0-4.0 mM beta-hydroxybutyrate.

**Exercise** both aerobic and strength training, was encouraged for at least 45 minutes per day, at least six days per week, and facilitated by the personal trainers. High-intensity interval training (HIIT) was recommended a minimum of twice per week.

**Sleep** hygiene was supported to ensure 7-8 hours of quality sleep per night, and all patients without known sleep apnea were tested over several nights using home sleep study devices. In those diagnosed with sleep apnea or upper airway resistance syndrome (UARS), referral for treatment with a continuous positive airway pressure apparatus (CPAP) or a dental splint device (for those identified with UARS) was provided.

**Stress** management included biofeedback and heart-rate variability training with a HeartMath Inner Balance for IOS device, for a minimum of 10 minutes per day.

**Brain training** was carried out using BrainHQ, a HIPAA and SOC-2-compliant platform with empirical validation^18^, for a minimum of 15 minutes daily. Participants trained on 29 cognitive exercises that target the speed and accuracy of information processing.

### Hormones and nutrients

For those patients with suboptimal hormonal status, bio-identical hormone replacement and appropriate supplements were provided to optimize sex hormone levels, neurosteroids, and thyroid medications as indicated for sub-optimal thyroid function. For those with suboptimal nutrients (e.g., vitamin D, omega-3, B vitamins, CoQ10, or minerals), the appropriate nutrients were provided.

### Gastrointestinal health

For those with gastrointestinal hyperpermeability, infections, inflammation, or impaired absorption and digestion, gut healing with dietary restriction, gut-healing nutrients, and digestive enzyme support if indicated, along with treatment of any identified dysbiosis, was undertaken.

### Inflammation

For those with evidence of systemic inflammation, pro-resolving mediators and anti-inflammatory herbal supplements were provided, low-dose naltrexone was prescribed (if there was evidence of autoimmunity), and omega-3 fats included via diet and supplements. Some patients utilized fasting-mimicking diets.

**Infectious agents** associated with cognitive decline or systemic inflammation were identified and treated. For those with evidence of *Herpes simplex* infection or a history of outbreaks, valacyclovir was prescribed for 2-6 months. Active *Epstein-Barr Virus* (EBV) was treated with herbal protocols. For those with evidence of tick-borne infections such as *Borrelia, Babesia*, or *Bartonella*, organism-sensitive treatment was prescribed with herbal anti-microbials and immune support.

### Toxins and toxicants

For those with toxicity associated with metals (e.g., mercury or lead), organic pollutants (e.g., benzene, phthalates, or organophosphate insecticides), or biotoxins (e.g., trichothecenes, ochratoxin A, or gliotoxin), targeted detoxification was undertaken with binding agents (e.g., cholestyramine or bentonite clay), sauna, herbs, sulforaphane, and dietary restriction of seafood if indicated.

## Results

### Cognition

The AQ-C is a subjective change scale that is derived from the AQ-21. It is informant-based (significant other or study partner) and has a range from −40 (marked decline in all functions) to +40 (marked improvement in all functions). A Likert-type scale was used, such that the scoring for each of the 20 questions was −2 (much worse), −1 (slightly worse), 0 (no change), +1 (slightly better), or +2 (much better).

Overall results were categorized as no change with a zero score, mild for a score change of 1 or 2 points (mild decline for −1 to −2, and mild improvement for +1 to +2), moderate for a change of 3 to 10 points, and marked change for 11 and greater. Table 2 displays the results.

**Table 2.** AQ-C informant-estimated changes.

| Result | Count | Percent |
| --- | --- | --- |
| Marked Decline (-11 to -40) | 2 | 8% |
| Moderate Decline (-3 to -10) | 1 | 4% |
| Mild Decline (-1 to -2) | 0 | 0% |
| No Change (0) | 1 | 4% |
| Mild Improvement (+1 to +2) | 3 | 12% |
| Moderate Improvement (+3 to +10) | 4 | 16% |
| Marked Improvement (+11 to +40) | 14 | 56% |
| **Total Improved** | **21/25** | **84%** |

Twenty-one of the 25 patients (84%) were rated as improved by their study partners. Using the Wilcoxon Signed Rank Test, the p-value for this outcome is 0.0005. Assuming a random distribution of estimates from study partners of improvement vs. decline, this result is also highly significant (p <0.01) by chi-square and binomial analyses. This indicates that the 25 participants overall experienced a subjective improvement (as judged by their study partners) over the course of the study period in which they received the treatment.

**CNS Vital Signs** is a computerized neuropsychological assessment used to evaluate cognitive performance and change. The reliability and validity of this battery have been described in previous publications,^19^ and this assessment tool is more sensitive than the MoCA in the identification of mild cognitive impairment.^20^ The test battery administered for this study included assessments of verbal memory, visual memory (both immediate and delayed), symbol digit coding, Stroop performance, shifting attention, continuous performance, and finger-tapping. Age-matched domain standard scores and percentile ranks were calculated for visual memory, verbal memory, composite memory, motor speed, psychomotor speed, processing speed, reaction time, cognitive flexibility, simple attention, complex attention, and executive function, as well as an omnibus domain score, Neurocognitive Index (NCI).

Figure 1 shows CNS Vital Signs results of the NCI from all patients who completed the study at baseline, 3 months, 6 months, and 9 months of treatment. Figure 2 summarizes medians and quartile results for the NCI at each time point. Comparing the results at outset to those at completion revealed an improvement in the Neurocognitive Index from 95.6 ± 8.4 to 105.0 ± 10.1 (p = 0.0001 by paired t-test), which corresponds to an increase from the 38^th^ percentile to the 63^rd^ percentile, and indicates an improvement that is highly statistically significant.

**Fig. 1.**
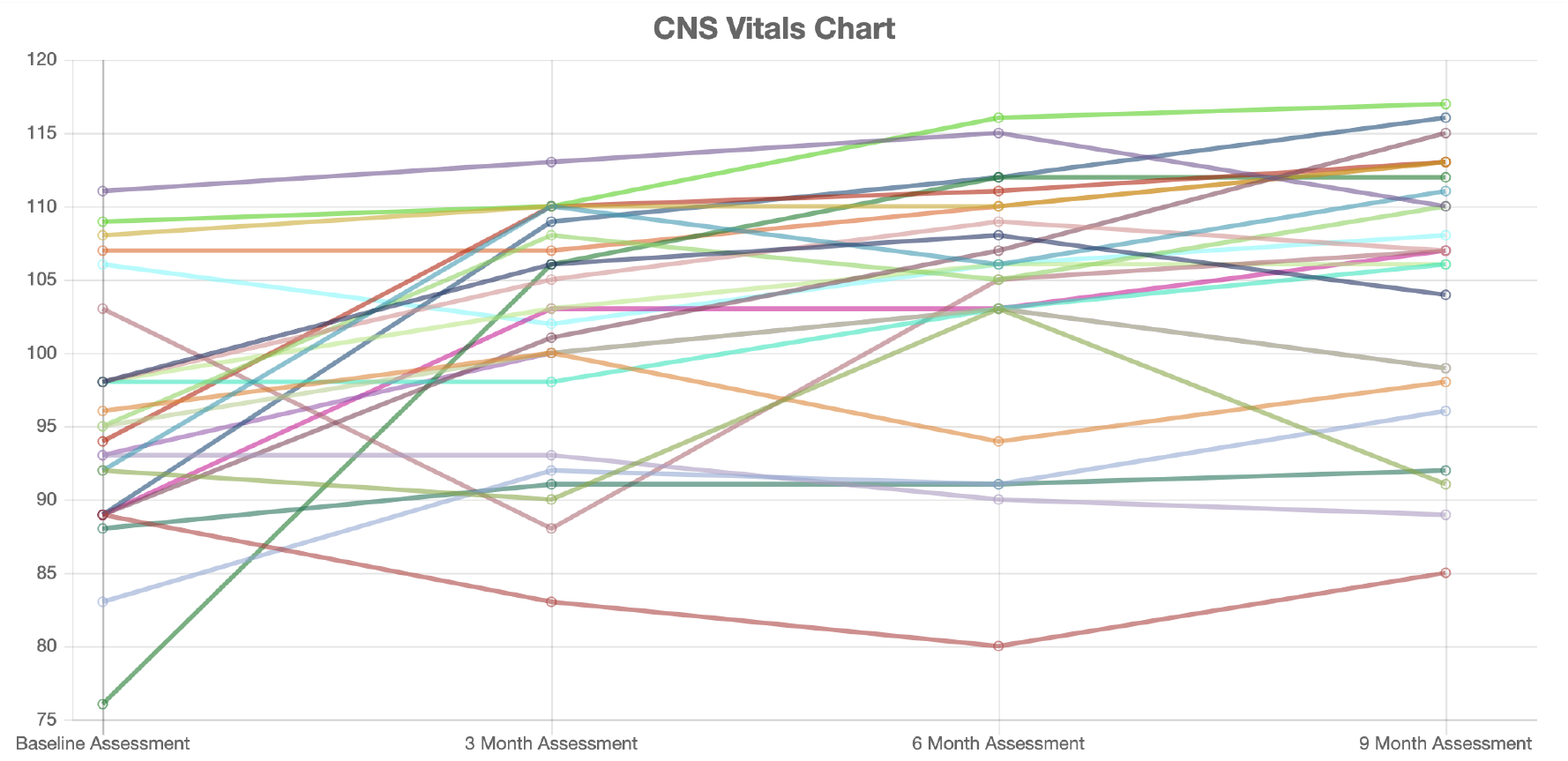
Neurocognitive index for all 25 patients at baseline, 3 months, 6 months, and 9 months of treatment.

**Fig. 2.**
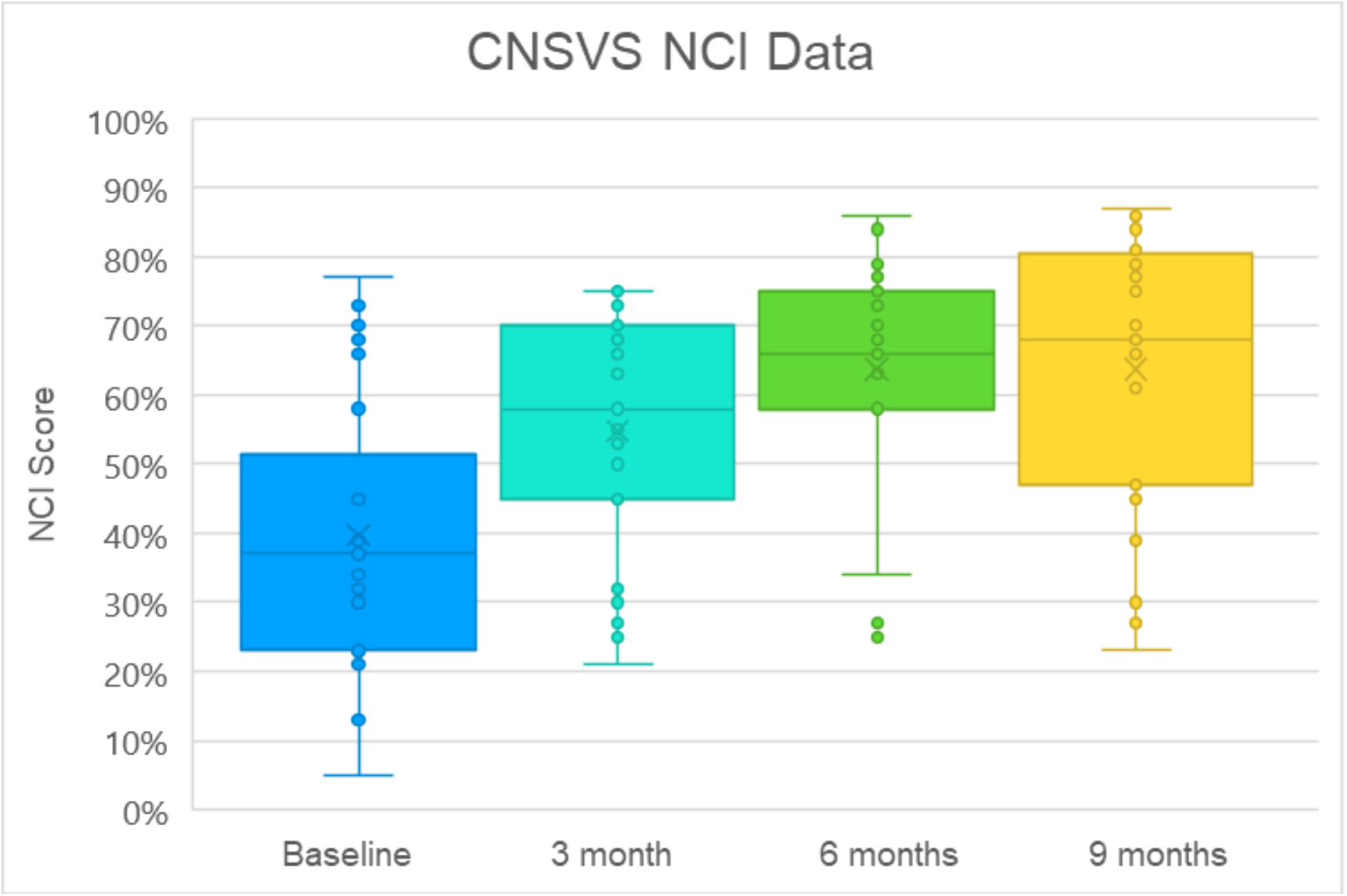
Neurocognitive indices (NCI) for the 25 patients at baseline, 3 months, 6 months, and 9 months. Dots represent scores, boxes represent second and third quartiles, horizontal lines within boxes represent medians, x represents means, and whiskers represent first and fourth quartiles. The ordinate represents NCI as percentile for age.

Visual analysis of the NCI data revealed it was not uniformly normal across time periods. As such, it was considered prudent to use a nonparametric related-samples Friedman’s two-way analysis of variance by rank to evaluate change across the four periods NCI data were collected (t=0, 3, 6, and 9 months). The overall test indicated a significant difference across the NCI time periods [chi square (3) 29.646, p<0.001]. Pair-wise comparisons with Bonferroni correction revealed that the baseline NCI score was significantly different from the NCI score at 3 months (p=0.023), 6 months (p<0.001), and 9 months (p<0.001), with NCI at 3 months significantly different from NCI at 9 months (p=0.006).

The range of percentile changes in the CNS Vital Signs NCI was from −9 (32^nd^ percentile → 23^rd^ percentile) to +74 (5^th^ percentile → 79^th^ percentile). Twenty-one of the 25 patients improved their CNS Vital Signs NCI scores (84%), one was unchanged (4%), two declined (8%), and one NCI (4%) was considered invalid due to visual field abnormalities.

Individual domains of the CNS Vital Signs battery were also evaluated. These domains were normally distributed on visual inspection and a repeated measures ANOVA was utilized. For tests that violated Mauchly’s test of sphericity, a Greenhouse-Geiser correction was utilized for reported p values. The repeated measures ANOVA revealed that mean CNS Vital Signs domain tests differed statistically significantly between time points, Pillai’s Trace 1.0 (*F*(10, 13) = 3001.696, *p* <0.001). Significant main effects were found for Psychomotor Speed (p<0.001), Executive Functioning (p<0.001), Motor Speed (p<0.001), Verbal Memory (p=0.007), Simple Attention (p=0.010), Cognitive Flexibility (p=0.013), and Reaction Time (p=0.036). All of these had significant linear trends indicating improved performance from baseline to 9 months except for Processing Speed, which displayed a significant quadratic trend, with the best performance being at 9 months.

**Montreal Cognitive Assessment** (MoCA) was also used to evaluate the patients (version 8.1 at baseline, 8.2 at 3 months, 8.3 at 6 months, and 8.1 at 9 months), and baseline MoCA scores ranged from 19 to 30, with a mean of 24.6 and standard deviation of 3.52. Final scores ranged from 19 to 30, with a mean of 27.56 and a standard deviation of 3.04 (Fig. 3).

**Fig. 3.**
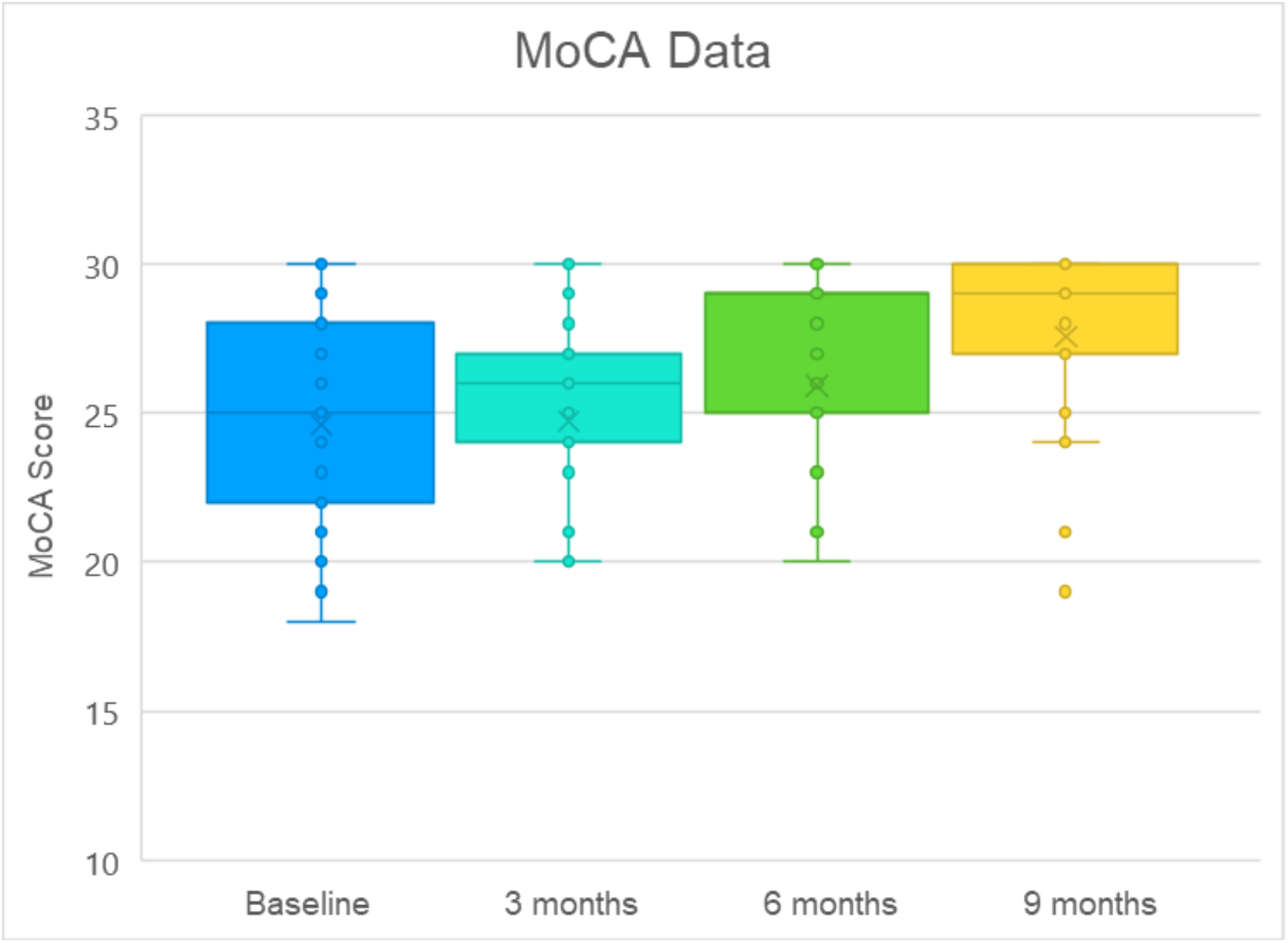
MoCA scores (dots), means (x), medians (bars; note that the median at 6 months was 29), and lowest and highest quartiles (whiskers) for the 25 subjects.

As the MoCA is not normally distributed and represents ordinal data, a nonparametric statistical test was utilized. A related-samples Friedman’s two-way analysis of variance by rank was used to evaluate change across the four periods during which MoCA data were collected. The overall test indicated a significant difference across the MoCA time periods [chi square (3) 16.923, p=0.001]. Pair-wise comparisons with Bonferroni correction revealed that the baseline MoCA score was significantly different from the MoCA score at 6 months (p=0.016) and 9 months (p<0.001), with MoCA at 3 months significantly different from MoCA at 9 months (p=0.006).

The range of MoCA changes was from −2 (21→19) to +11 (19→30). Of the 25 patients, 19 (76%) improved their scores, 3 (12%) showed decline in scoring, and 3 (12%) were unchanged. These results are compatible with those from the AQ-C and CNS Vital Signs in demonstrating improvement that is highly significant statistically. For comparison, historical controls without cognitive complaints reduced their MoCA scores by an average of 0.52 points annually.^21^

**Brain training** was, as noted above, undertaken by all patients, and the performance on BrainHQ, although intended as an interventional component, offers preliminary insight into cognitive performance and its decline or improvement during the 9-month trial. All 25 of the patients improved their BrainHQ percentile composites during the trial, with a mean improvement of 21% (range: 9% - 33%).

### Brain MRI with volumetric quantification

Gray matter volumes of the trial patients were increased by a mean of 0.3% on an annualized basis. For comparison, longitudinal gray matter volumes typically decrease by an average of 0.83-0.92% per year for those without cognitive decline^22^, and 2.20-2.37% for those with Alzheimer’s^23^.

Hippocampal volumes of the trial patients were decreased at an annualized rate of 1.29%. For comparison, hippocampal volumes decrease in patients with MCI or Alzheimer’s disease at an annualized rate of 3.5-4.66%, and in cognitively stable controls at an average rate of 1.41-1.73%.^24,25^ (Note, however, that those references did not utilize the same computerized analysis of gray matter and hippocampal volumes used here, which may or may not have impacted results.) Thus both gray matter volume changes and hippocampal volume changes were observed to be better than expected—not only in comparison to patients with Alzheimer’s but also in comparison to healthy aging adults—based on historical studies.

## Discussion

The results of this proof-of-concept trial support the performance of a larger, randomized, controlled clinical trial. The magnitudes of effects, proportion of patients improved, and combinations of improvements observed here—in MoCA scores, CNS Vital Signs scores, AQ-C, BrainHQ, and MRI volumetrics—have not been reported previously. Thus the overall results support the notion that a precision medicine approach to the cognitive decline of Alzheimer’s disease and mild cognitive impairment may be an effective strategy, especially with continued optimization over time.

The approach utilized in this trial departs sharply from traditional treatment strategies for MCI and Alzheimer’s disease, which have largely been monotherapeutic, monophasic, non-personalized, and blind, i.e., cause-independent, thus not targeted to the underlying drivers of the disease in each person, but rather to common downstream consequences and/or secondary drivers, such as amyloidosis. This is likely at least in part because Alzheimer’s disease remains a disease of unknown, and controversial, etiology, with many competing theories, none of which has led to effective treatment. The dominant theory over the past three decades has been the amyloid cascade hypothesis^26^, but numerous antibodies targeting the associated amyloid have failed to improve cognition (although a recent trial that failed to improve cognition or halt decline nevertheless slowed decline by 32%^27^).

The strategy utilized in this study also differs from preventive management of Alzheimer’s disease risk factors ^28^, an emerging strategy whose interventions are informed by statistical associations rather than by individual network diagnostics, and whose main purpose is to delay rather than to halt and reverse cognitive decline, although both strategies are compatible in ideology and complementary in practice.

The positive results from the proof-of-concept trial reported here are compatible with the notion that Alzheimer’s disease represents a complex network insufficiency. Therefore, multi-factorial optimization of network function and support offers a rational therapeutic strategy. It might be argued that the strategy utilized for this trial targets associated biochemical pathways but not necessarily causal ones. However, both the utilization of genomics to help identify underlying causal factors and the cognitive improvements documented in this study argue that at least some of the biochemical targets addressed are indeed causal. Moreover, due to the small-world nature of biochemical and signaling networks, targeting a sufficient number of associated pathways is likely to impact the causal ones, even if indirectly. Nonetheless, it will be important for future studies to continue to dissect and prioritize the targeted interventions in order to develop an optimal protocol for each individual.

One potential concern regarding the positive results reported here is whether they may simply be due to practice effects. There are several points that make this possibility highly unlikely: (i) the CNS Vital Signs testing has been designed to minimize such effects, and this has been demonstrated experimentally^29^; (ii) the 3-month interval renders practice effects less likely than shorter duration intervals; (iii) the magnitude of the effects (e.g., MoCA increases from 19→30) are incompatible with practice effects, which are typically much more modest; (iv) different MoCA tests were used for baseline, 3-month, and 6-month evaluations, with 9-month repeating the baseline version, in order to minimize practice effects; (v) the AQ-C score improvements provided confirmation of the increased MoCA and NCI scores.

There are several points worthy of further clarification and discussion: first, potential trial patients with MoCA scores of 18 and lower were excluded from this study, and therefore, although there are anecdotal reports of patients with such scores showing improvement with a similar precision medicine approach,^30^ the current study offers no insight into the treatment of patients in that group.

Second, the MoCA score improvements reported here suffered from a ceiling effect, because several patients with MCI who met the entry criteria based on cognitive complaints, AQ-21 >5, and qualifying CNS Vital Signs scores had MoCA scores of 28-30 at baseline. With the inclusion of those 7 patients, the mean change in MoCA score was +2.96, whereas removing those patients increased the mean MoCA change to +3.89. Nonetheless, even with the inclusion of those patients, the improvement in MoCA scores was highly significant. Furthermore, the more sensitive CNS Vital Signs test^31^ provided a dynamic range for these 7 patients with baseline MoCA of 28-30 (despite clear cognitive complaints and AQ-21 >5), and 6 of the 7 improved their NCI, with the other remaining unchanged.

Third, the COVID-19 pandemic began during this trial, and may have impacted some of the results. Many of the patients found themselves unable to visit gyms and swimming pools, unable to work with personal trainers or health coaches, sheltering in place in homes with high mycotoxin levels, having more difficulty obtaining the food necessary for the diet used in the protocol (many elderly were encouraged not to leave home early in the pandemic), and with increasing social isolation. A few of the patients reduced their compliance in association with the pandemic instructions for sheltering in place and social distancing. In 7 of the 25 patients, their 6-month MoCA scores were slightly higher than their 9-month scores, and similar effects were seen for the NCI scores. Thus, without the pandemic, the improvements documented here may have been greater. However, although the long-term effects may be more substantial, the overall effects on the scores did not affect the high degree of statistical significance.

Fourth, the MRI effects observed, although promising, were modest. The gray matter volume change of +0.3% is ostensibly different than the expected −2.20-2.37%, but further studies will be required to confirm that gray matter atrophy is affected by the approach used here. Similarly, the hippocampal volume change of −1.29% observed is better than the −3.5% to −4.66% expected from previous studies on patients with MCI or dementia,^32^ and better than even the more modest loss (−1.73%) recorded in cognitively stable controls, yet further studies will be required to confirm this effect.

Fifth, a previously reported precision medicine approach to cognitive decline^33^ showed no significant improvement in those with MCI or dementia, in contrast to the results reported here. However, that study was more modest in both the evaluation and treatment protocols employed—for example, many of the pathogens and toxins evaluated and treated in the current study were not addressed in that study—and therefore, it is possible that success with such an approach will require identifying and targeting the many potential contributors to cognitive decline, as opposed to restricting the therapy to a more limited subset.

Sixth, this proof-of-concept trial lacks a direct comparison to patients treated pharmacologically or untreated. However, historical data have shown repeatedly that patients with MCI or early dementia undergo a downhill course;^34^ moreover, the trial noted above^35^ included analysis of low-compliance patients with MCI or early dementia, with initial deficits similar to the group reported here, and recorded decline in those patients. Therefore, our finding of improvement relative to baseline would likely reveal even greater improvement if compared to a control group, since decline rather than stability is the observed natural course of MCI and early dementia.

Seventh, there were no serious adverse events recorded in this study. On the contrary, most patients improved their overall health, and unpublished observations show that some patients will no longer require anti-hypertensives, anti-diabetes drugs, or lipid-lowering agents, as they address the contributors to cognitive decline. This is compatible with the approach of identifying and targeting the root cause contributors to cognitive decline, improving resilience and overall health.

Eighth, since cerebrospinal fluid was not analyzed for amyloid-beta peptide and tau in these patients, it may be argued that they did not have Alzheimer-associated pathology. However, the vast majority of patients in this age group who show progressive cognitive decline, with AQ-21 >5, abnormal MoCA scores, and who are ApoE4+, do indeed have Alzheimer-associated pathology. If the protocol used here were only effective for patients with non-Alzheimer’s-associated pathology, then the ApoE4+ group would be expected to do worse than the ApoE4-negative group. However, both groups—the ApoE4+ group and the ApoE4-group—showed statistically significant improvements in MoCA and NCI. This does not exclude the possibility that some of the patients in the study could have had non-Alzheimer’s pathology, but it supports the conclusion that the protocol used is effective for patients with Alzheimer’s pathophysiology, at least those with MoCA scores of 19 and higher.

Finally, this study confirms and extends anecdotal reports that it is possible to reverse cognitive decline in MCI and early dementia with a personalized, precision medicine protocol, but it does not show that it is practical to do so. The analysis involved is more comprehensive than is currently in use in memory centers, the data sets collected more extensive, the behavioral alterations required of the patients more demanding, the time required by the team of practitioners greater, and the cost significant (although far less than an assisted living facility). Further refinement and simplification of the protocol may render it more feasible, accessible, and affordable. Furthermore, given the recognized biochemical targets of the interventions, novel pharmaceutical agents may become a critical part of an optimal protocol, and, in a complementary fashion, future trials of new drug candidates may enjoy more successful outcomes when conducted in the context of precision medicine protocols.

## Data Availability

All data (de-identified) in the manuscript are publicly available.

## Acknowledgments

We are grateful to the Four Winds Foundation for its support, to Diana Merriam and Gayle Brown for their support of this study, and to Dr. David Perlmutter for introductions. For providing diagnostics for the trial, we thank CNS Vital Signs, Posit Science, HeartMath, Great Plains Laboratory, Cyrex Laboratory, Armin Labs, Bio-Botanical Research, Genova Diagnostics, Doctors Data, IntellxxDNA, RadNet, and Norcal Imaging. We also thank Brainreader ApS for providing use of the Neuroreader software for the imaging analyses in this study. For clinical support, we thank Michael Atkinson, Derek Barber, Mac Dodds, James Gaydos, Doug Jaser, Lynn Killips, Karen Preskenis, Renée Riley-Adams, Venessa Rodriguez, Kia Sanford, and Sheila Wagner. For research support leading to this trial, we are grateful to Phyllis and James Easton. We thank Dr. Alexei Kurakin and Dr. Aida Lasheen Bredesen for comments on the manuscript. The funders of this study had no role in study design, data collection, data analysis, data interpretation, or writing the report. Dr. D. Bredesen consults for Apollo Heath, which had no role in study design, data collection, data analysis, data interpretation, or writing the report.

